# Is the fit of N95 facial masks effected by disinfection? A study of heat and UV disinfection methods using the OSHA protocol fit test

**DOI:** 10.1101/2020.04.14.20062810

**Authors:** Amy Price, Yi Cui, Lei Liao, Wang Xiao, Xuanze Yu, Haotian Wang, Mervin Zhao, Qiqi Wang, Steven Chu, Larry Chu

## Abstract

The current COVID-19 pandemic has highlighted global supply chain shortcomings in the US hospital delivery system, most notably personal protective equipment (PPE) and COVID-19 is found on these masks ∼7 days. Recent work from our group has shown two promising disinfection methods for N95 facial masks, dry heat (hot air (75 °C, 30 min) and UVGI which is UVGI 254 nm, 8W, 30 min. Using N95 five models of N95 masks from three different manufacturers we determined the following: 1) Hot air treated N95 masks applied over 5 cycles did not degrade the fit of masks (1.5% change in fit factor, p = .67), 2) UVGI treated N95 masks applied over 10 cycles were significantly degraded in fit and did not pass quantitative fit testing using OSHA testing protocols on a human model (−77.4% change in fit factor, p = .0002).

**NOTE:** *We would like to share our results with the community as soon as possible. Be mindful that this report is a pilot study and a work in progress. We will have more results in the coming days and weeks*.

We recommend that **hospital policy** and **procedures** be respected and adhered to. Do not use anything in your home to disinfect contaminated equipment. Please do not heat your masks in a home oven!

## Introduction

The current COVID-19 pandemic has highlighted global supply chain shortcomings in the US hospital delivery system, most notably personal protective equipment (PPE). Frontline health care workers across the United States report shortages of PPE ranging from gloves, protective gowns, eye wear and face masks.

The transmission of COVID-19 is thought to occur through respiratory droplets, and current CDC guidelines recommend the use of N95 masks for health care providers managing the care of patients infected with SARS-CoV-2 or persons under investigation (PUI) for COVID-19.

The global shortage of PPE in the setting of a viral pandemic has created potentially dangerous conditions for frontline healthcare workers lacking appropriate protection and their patients. This has led to recent efforts to extend existing PPE resources. Suggested solutions such as extended wear Bergman et al.^1^, frequent donning and doffing Brady et al. ^2^and the appearance of the virus on N95respirator masks for ∼7 days Chin et al^3^ increase risk to the population and specifically to healthcare providers.

Many hospitals have initiated efforts to treat N95 respirators to deactivate the SARS-CoV-2 virus so they may be reused by health care workers. The CDC has recently released guidelines on extended use and limited reuse of disposable N95 respirators in the health care setting. In their guidance, the CDC specifically cautions that “A key consideration for safe extended use is that the respirator must maintain its fit and function.” However, there is a dearth of high-quality evidence evaluating fit and function of masks after such treatments.

The CDC recommends that <u>PPE be provided to health care professionals</u> according to OSHA PPE Standards, including proper cleaning and disposal.^4^ It is also mandatory that face masks be fit tested using <u>OSHA-accepted fit-testing protocols</u> prior to use.^5^ OSHA does not currently have PPE standards for disinfecting N95 masks.

<u>OSHA states</u>, “The N95 filtering facepiece respirator is a “disposable respirator.” It must be discarded after use, or when it becomes damaged or soiled. It cannot be cleaned and disinfected according to Appendix B-2. OSHA is presently not aware of any alternate procedures provided by respirator manufacturers in their user instructions that would allow for cleaning and disinfecting their filtering facepiece respirators.”^6^

The largest N95 respirator manufacturer 3M admonishes that disinfection be effective against the target organism, such as the virus that causes COVID-19, retain respirator filtration, not affect the respirator’s fit; and be safe for the person wearing the respirator. If the filtration is damaged or the respirator does not fit, it will not help reduce exposure to airborne particles at the level indicated, such as N95. They report on March 27,2020, no disinfection method met all four of these key criteria, and without all four, the method is not acceptable^7^.

Masks that have been heat treated, UVGI treated or repeatedly donned and doffed^8^may lose their shape and require fit-testing, as we stated in our previous Evidence Service report. The CDC recommends limiting how germs enter a facility (or home)^4^. Likewise, materials consistently donned and doffed have potential for contamination.

In this study, we are testing the materials used by 3 mask manufacturers (3M, Jackson, and 4C) for filtration and fit. No one that we know of has yet tested these materials with SARS-CoV-2 virus as the use is tightly controlled and limited to level 3 labs.^9^ All disinfection methods need to be tested.^10^ At this point known pathogens with similar size are used to test for filtration and to estimate inactivation efficacy.

## RATIONALE FOR FIT

A respirator fit test checks that there is an airtight seal between the wearer’s respiratory system and the ambient air. This is achieved by a mask fit that is conforms to the face (without gaps) to ensure a seal on the mask perimeter. Because wearers are different sizes and shapes and are not protected if there are gaps, it is necessary to test the fit of the mask.

Work by others have evaluated the fit and function of masks after various disinfection treatments. These fit testing methods have utilized mannequin heads, and have used a small number of fit tests (usually 3) compared to the OSHA testing protocol for human fit testing (8 tests). This is due to the fact that mannequin heads do not move. Humans model fit testing involves 8 tests, including: 1) normal breathing, 2) deep breathing, 3) turning the head left-to-right, 4) moving the head up-and-down, 5) talking, 6) grimacing, 7) bending over, and 8) normal breathing. Therefore, to our knowledge, there have not been any studies that have measured quantitative fit testing on heat- and UVGI-disinfected treated masks using a human model on the OSHA protocol. This would be the most accurate method to predict the fit performance of a mask on health care workers after disinfection. As authors of previous mannequin-based studies have stated as limitations of their studies, “These results should not be interpreted as being equivalent to a standard OSHA-accepted fit test method^11^ Human subject testing is needed to truly assess respirator fit.”

We will first outline the decontamination methods described in detail elsewhere and then go on to describe the methods for mask fit.^12^

## Methods

### IRB EXEMPTION

Based on the information provided about “Does reuse and or disinfection of N95 masks effect fit”, the Research Compliance Office, Stanford University determined on March 23, 2020 that this research does not involve human subjects as defined in 45 CFR 46 nor 21 CFR 56.

### DISINFECTION METHOD 1: HOT AIR (75 °C)

1. Preheat the oven (Figure 2) to 75 °C.

**Figure 1:**
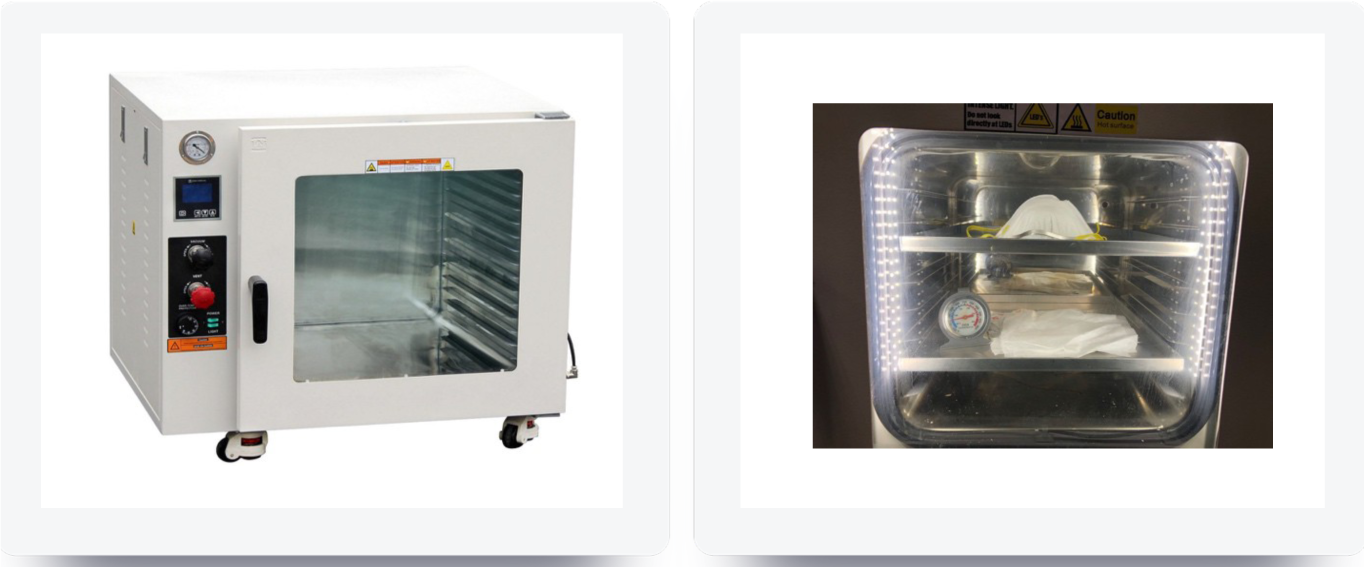
AI 14 Shelf Max 5 Cu Ft 480°F 5-Sided Heating Vacuum Oven, Across International LLC.

**Figure 2:**
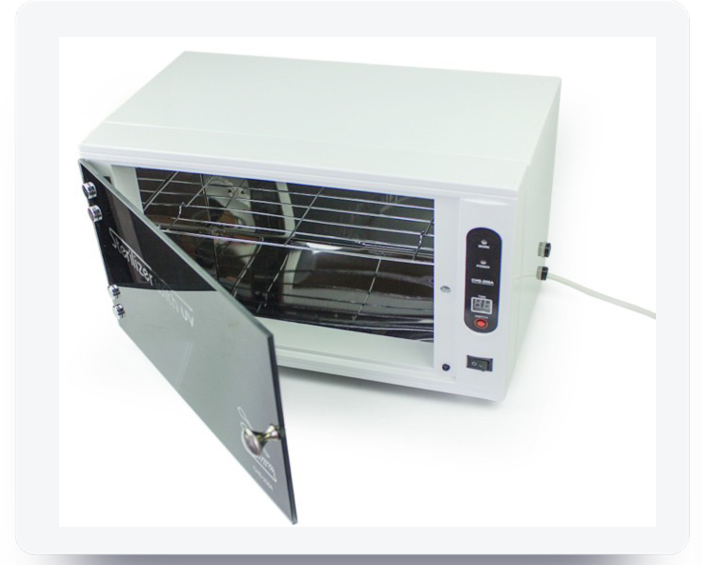
Sterilizer Cabinet With UV, CHS-208A, 110V/60 Hz, 254 nm
2. Put the N95 masks to be tested into the oven.
3. After 30 minutes of heating, take out all the samples and cool at room temperature for 10
4. minutes.
5. Repeat Step 3 for a total of 5 total cycles.

### DISINFECTION METHOD 2: UVGI

1. Place samples into a UV sterilizer (Figure 3, 254 nm wavelength, 8 W UV light bulb).

**Figure 3:**
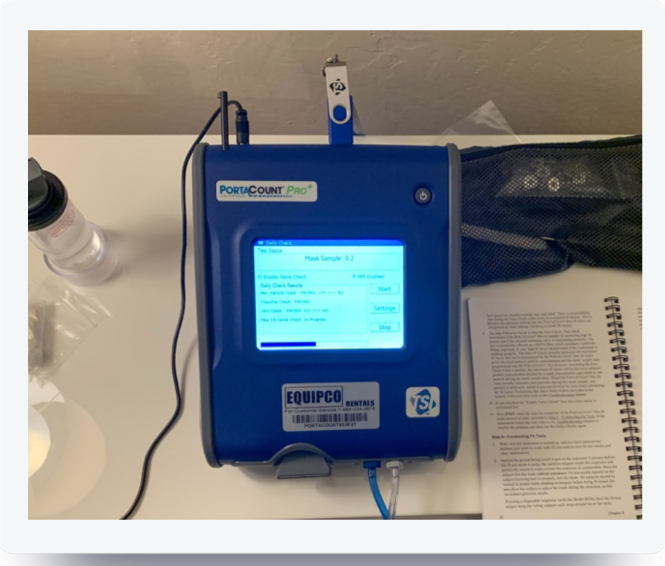
Fit testing data for 4C AIR N95 masks. Data for fit testing were obtained using the OSHA Protocol on a human model with the Portacount Pro+ 8038 respiratory fit tester. Daily quality control checks were performed prior to testing.
2. Irradiate under UV light for 30 minutes.
3. Take out the samples and let stand under ambient conditions for 10 minutes.
4. Repeat Steps 1-2, for a total of 10 cycles.

### FIT TESTING OF N95 RESPIRATORS

Fit testing was done with the PortaCount® Pro+ 8038, (Shorewood MN). A quantitative respirator fit test system approved for testing disposable filtering facepieces Series 100, 99, P3, and HEPA masks, half and full-face tight-fitting respirators, gas masks, PAPR, and SCBA masks, plus N-95, P2, and P1 disposables. Fit testing of masks was done with OSHA approved protocols for testing N95 disposable masks (plus P1 and P2).

1. The Portacount Pro + 8038 respiratory fit machine was first calibrated for testing using internal quality assurance checks, including particle check, classifier check, valve check, zero check, and max fit factor check. Testing did not proceed unless the machine passed all quality control checks (Figure 3).
2. A test probe was inserted into disposable N95 respirators according to the make and model of the respirator. For proper fit testing, the probe was inserted into the “breathing zone” of the respirator where it would not be blocked by the person’s nose or chin (Figure 4). For the KN95 respirator (4C Air, Sunnyvale, CA) this was located to the left of the fold and for the 3M 8210 Plus respirator (3M, St. Paul, Minnesota) this was located above the fold (Figure 5–6).

**Figure 4:**
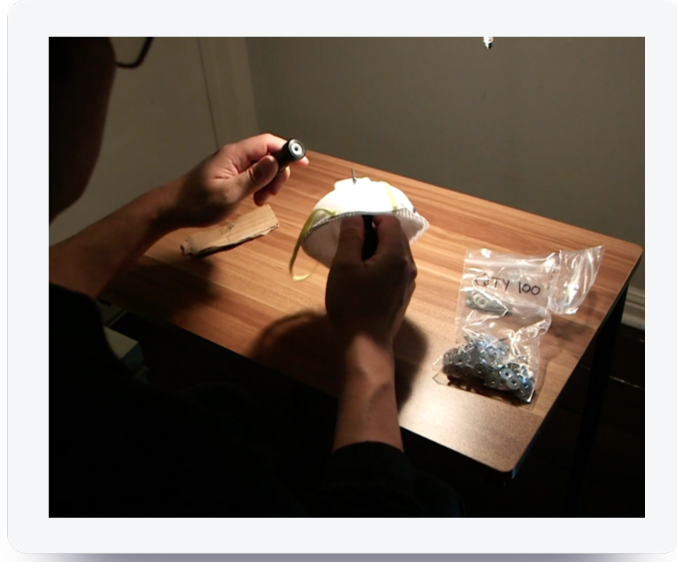
Sample probes were placed in disposable N95 masks in the “breathing zone” of the respirator according to the specific make and model of the mask so that the probe would not be blocked by the person’s chin or nose. The model is a co-author of this paper (LC)

**Figure 5:**
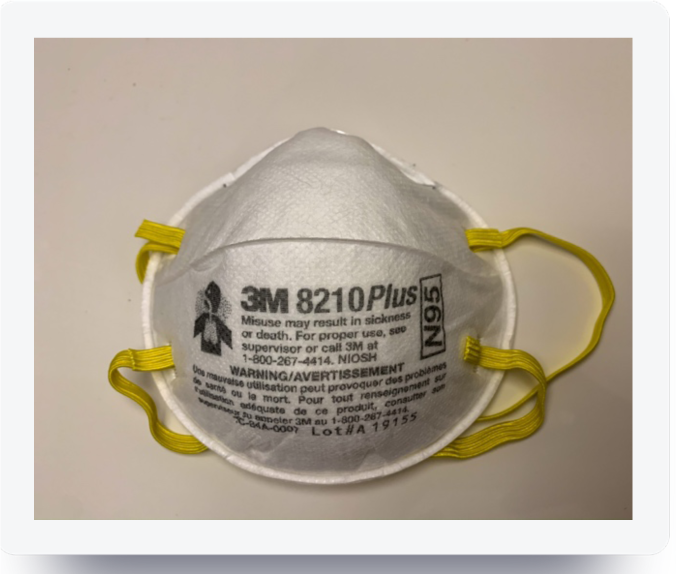
The 3M 8210 Plus N95 respirator prior to sample probe placement.
3. Fit testing of test masks was performed on a human model (LC) using the OSHA test protocol of the Portacount Pro+ 8038. This consisted of eight dynamic tasks: normal breathing (Figure 7), heavy breathing, turning head side-to-side, head up-and-down (Figure 8), talking, grimace, bend over and normal breathing. Each cycle of tests was performed 2 times for each mask.

**Figure 6:**
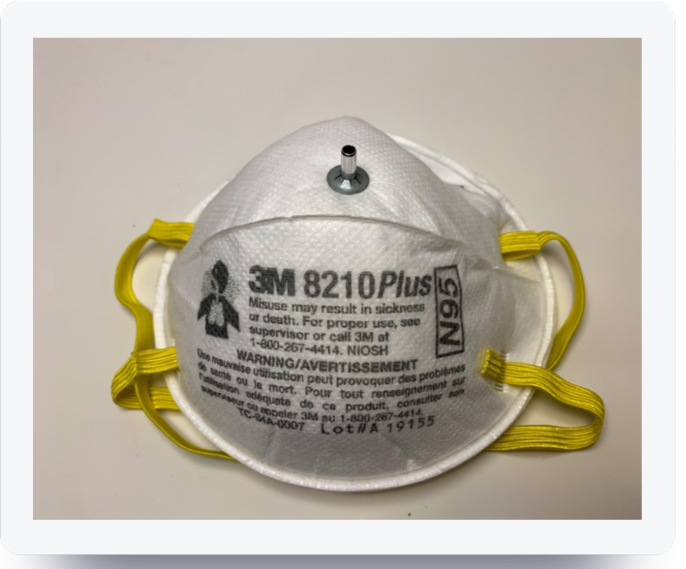
The 3M 8210 Plus N95 respirator after sample probe placement.

**Figure 7:**
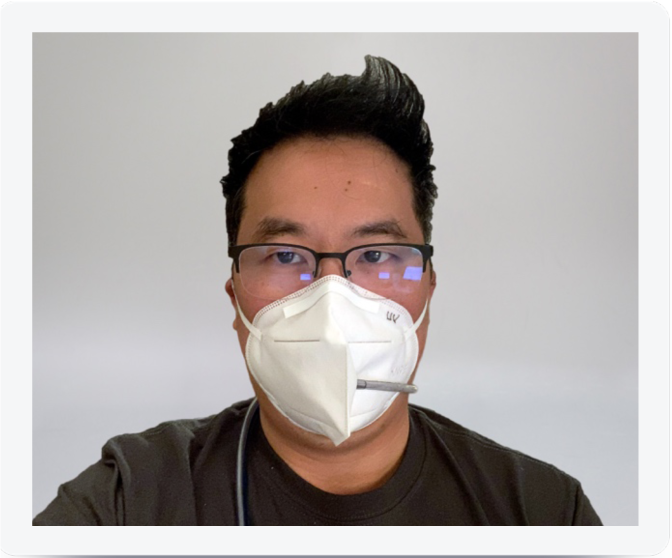
The OSHA test protocol was performed on a human model and uses dynamic tests. Normal breathing is demonstrated here. The model is a co-author of this paper (LC)

**Figure 8:**
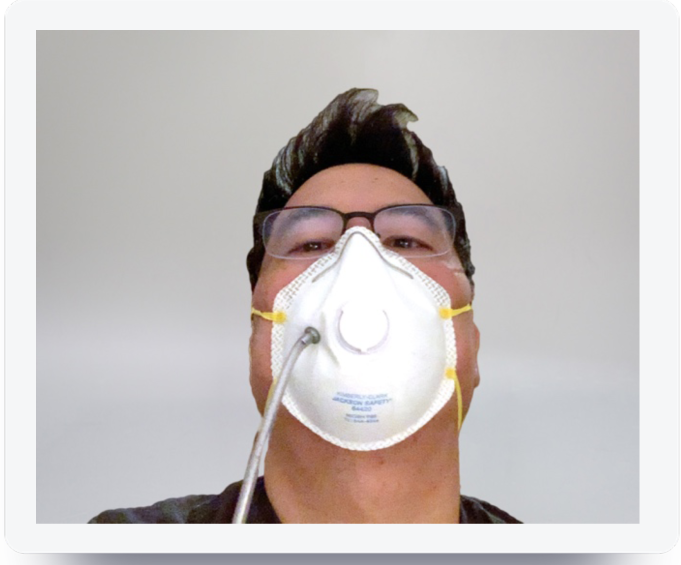
The OSHA test protocol was performed on a human model and uses dynamic tests. Head up-and-down is demonstrated here.The model is a co-author of this paper (LC)
4. Fit factor is computed as follows; 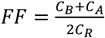 where:
  a. FF = fit factor
  b. C_B_ = particle concentration in the ambient sample before the respirator sample
  c. C_A_ = particle concentration in the ambient sample after the respirator sample
  d. C_R_ = particle concentration in the respirator sample
  e. The fit factor for individual component dynamic tests was computed for each task (e.g. normal breathing, deep breathing, etc.)
5. Overall fit factor is computed as follows; 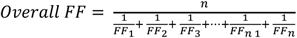 where:
  a. FF_x_ = fit factor for test cycle
  b. n = number of test cycles (exercises)

## STATISTICAL ANALYSIS

In the present study, the data analysis was conducted in two steps: descriptive statistical analyses and main hypothesis testing analyses. The descriptive statistical analyses computed the main outcome measures by treatment group. These analyses consisted of a series of univariate measures of central tendency computed to summarize continuous data using mean. Measures of dispersion were computed to understand the variability and were computed using standard deviation. The main analysis was tested using a series of hypotheses by univariate approach. Association between treatment method and overall fit factor was tested using a one-way ANOVA procedure (α=0.05). The final set of analyses used independent sample t-tests (α=0.05) to test the individual post-hoc associations between treatment types for significance, using the Benjamini-Hochberg procedure (false discovery rate of 0.25) to control for multiple comparisons. We report raw uncorrected p-values in this study and state a result is significant if the p < 0.05 after the Benjamini-Hochberg procedure. Statistical software SPSS for Mac 26.0 (Armonk, New York) was used to compute analysis for this study.

## Results

### FIT TESTING DESCRIPTIVE STATISTICS

We found that heat treatment did not significantly decrease the overall fit factor of KN95 masks (− 0.56% change) or 3M 8210 masks (0% change) compared to untreated control masks. We did find however that UV treated KN95 masks had, on average, a 90% decrease in overall fit factor compared to untreated controls.

**Table 1:** Fit factor of five models of N95 respirator masks without treatment, after hot air treatment, and after UVGI treatment. The 3M 8210 mask was not treated with UVGI due to a shortage of testing samples. The 3M 8200 mask was not tested due to broken straps.

| By Treatment Group |  |  |  |
| --- | --- | --- | --- |
| GROUPS | COUNT | MEAN (STD) | % CHANGE |
| Control (overall) | 10 | 191.95 ± 17.04 | – |
| Heat (overall) | 10 | 194.75 ± 11.81 | 1.5% |
| UVGI (overall) | 8 | 43.3 ± 65.44 | -77.4% |
| By Manufacturer |  |  |  |
| GROUPS | COUNT | MEAN (STD) | % CHANGE |
| 4C Air KN95 Control | 2 | 178.25 ± 43.5 | – |
| 4C Air KN95 Heat | 2 | 176.25 ± 33.78 | -0.56% |
| 4C Air KN95 UVGI | 4 | 17.45 ± 5.90 | -90% |
| 3M 8210 Plus N95 Control | 2 | 200 ± 0 | – |
| 3M 8210 Plus N95 Heat | 2 | 200 ± 0 | 0% |
| 3M 8210 Plus N95 UVGI | 0 | N/A | N/A |
| 3M 8200 Control | 2 | 181.5 ± 26.16 | – |
| 3M 8200 Heat | 2 | 200 ± 0 | 10.1% |
| 3M 8200 UVGI | 2 | Mask fail/straps broken | N/A |
| 3M 8511 Control | 2 | 200 ± 0 | – |
| 3M 8511 Heat | 2 | 200 ± 0 | 0% |
| 3M 8511 UVGI | 2 | 129.5 ± 99.70 | -35% |
| Jackson R20 Control | 2 | 200 ± 0 | – |
| Jackson R20 Heat | 2 | 197.5 ± 3.54 | -1.25% |
| Jackson R20 UVGI | 2 | 8.8 ± 4.53 | -95.6% |

\* % change in fit factor compared to untreated control samples for manufacturer's mask.

**Figure 9:**
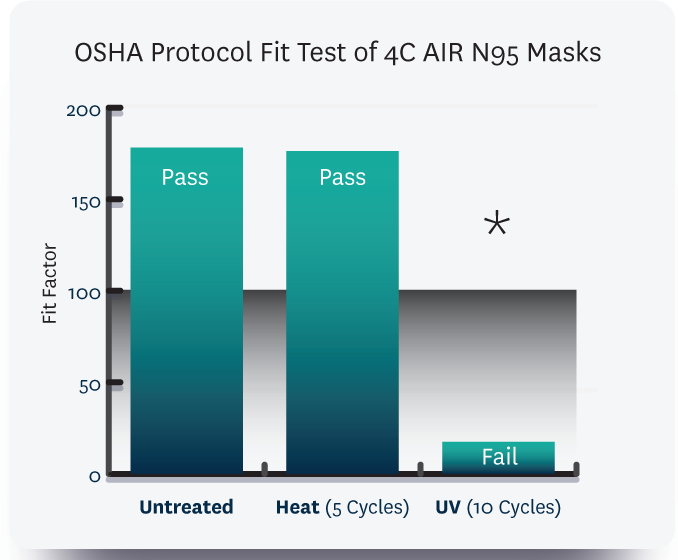
Fit testing data for 4C AIR N95 masks. Data show a that heat does not significantly decrease fit factor, however UVGI treatments decrease fit factor by 90% (p = .02), which was statistically significant.

**Figure 10:**
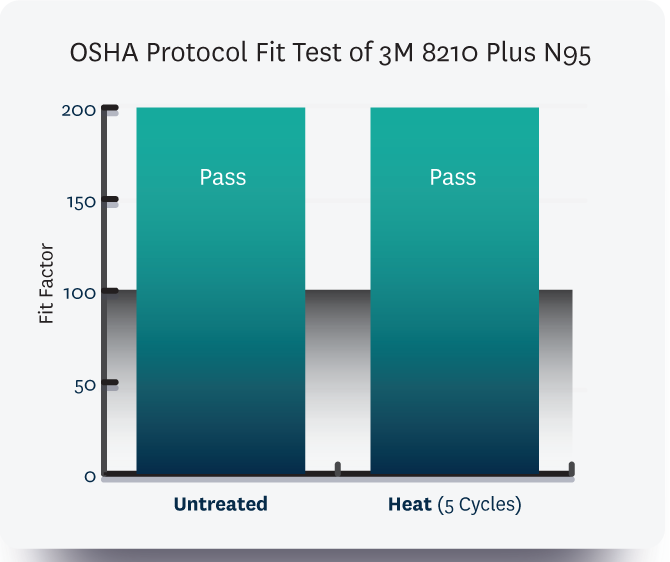
Fit testing data for 3M 8210 Plus N95 masks. Heat treatment did not significantly effect fit factor for these masks.

**Figure 11:**
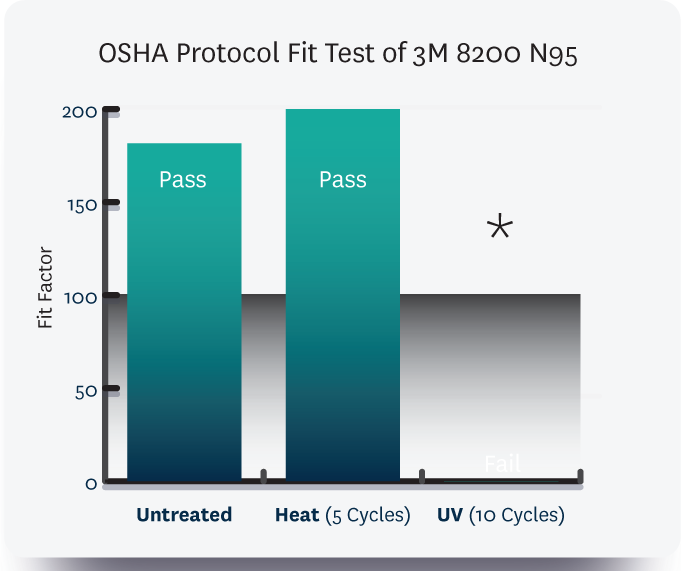
Fit testing data for 3M 8200 N95 masks. Data show a that heat does not significantly decrease fit factor, however UVGI treatments cause both masks to fail fit testing due to broken straps (prior to fit testing).

**Figure 12:**
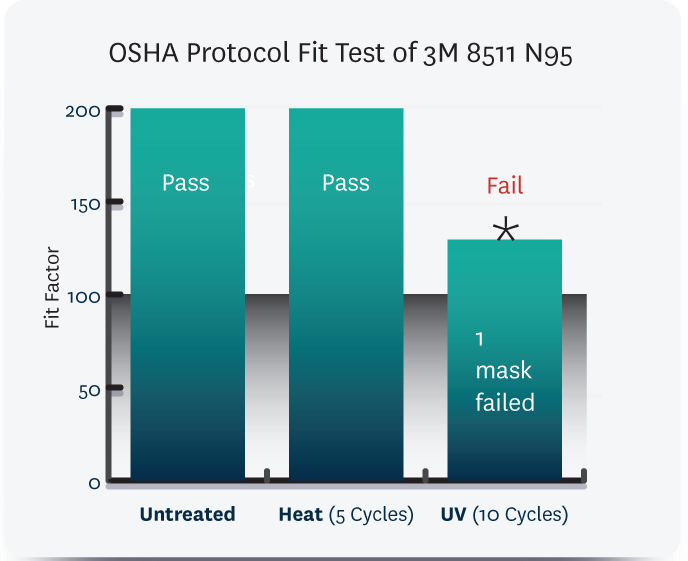
Fit testing data for 3M 8511 N95 masks. Heat treatment did not significantly effect fit factor for these masks, 1 mask failed fit testing after UVGI treatment (−35%, p = .42).

**Figure 13:**
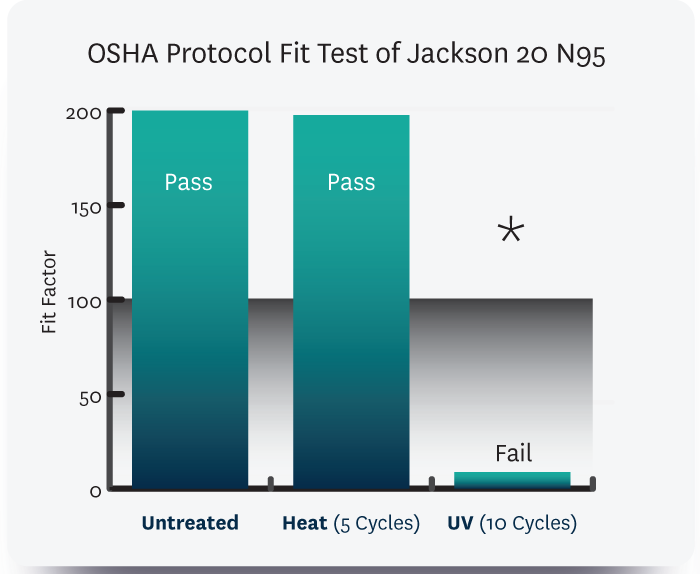
Fit testing data for Jackson 20 N95 masks. Data show a that heat does not significantly decrease fit factor, however UVGI treatments decrease fit factor by 95.6% (p = .0000028), which was statistically significant.

**Figure 14:**
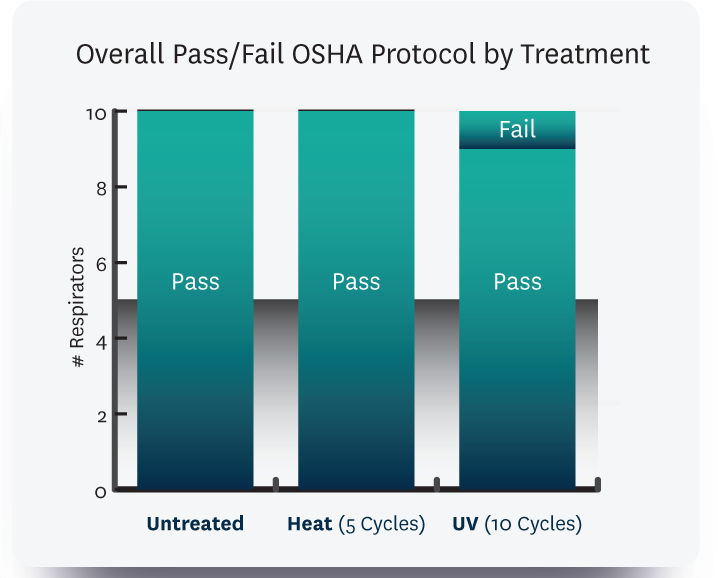
Overall Pass/Fail for 3 manufacturers of N95 masks. Data show that heat does not significantly decrease fit factor, however UVGI treatments decrease fit factor on average 77.4% (p = .00024), which was statistically significant.

**Figure 15:**
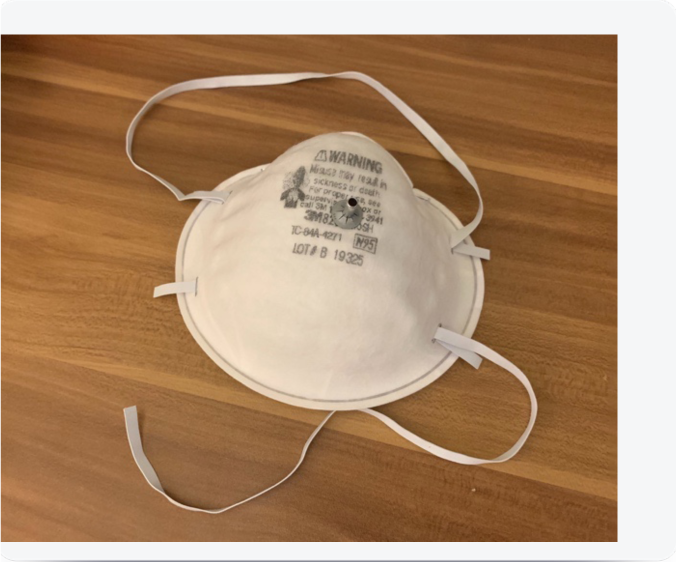
The 8200 mask removed from UVGI treatment prior to placement on human model. Straps were not manipulated or stretched. The lower strap was noted to be broken on both UV-treated masks.

## HYPOTHESIS TESTING

Our H_0_ (null hypothesis) = Disinfection treatment of masks (heat or UV) will not change the fit factor of masks as measured using OSHA standard protocols with a human model on the quantitative fit tester. To demonstrate effectiveness (preservation of fit), the treatment would show little or no change between the control (no treatment) and experimental (75°C Hot Air (30 mins) for 10 cycles) and UVGI (254 nm, 8W, 30 min) for 10 cycles) conditions.

### TREATMENT GROUP EFFECTS

There was no significant effect between heat treatment for the ten heat-treated masks (M = 194.75, SD = 11.81) in comparison to the ten untreated masks in the control group (M = 191.95, SD = 17.04) on fit factor scores t(18) = −0.4271, p = .67.

There was a significant effect between UV treatment for the ten UV-treated masks (M = 43.3, SD = 65.44) in comparison to the ten untreated masks in the control group (M = 191.95, SD = 17.04) on fit factor scores t(8) = 6.2572, p =.00024.

### MASK MODEL GROUP EFFECTS

There was a significant effect of treatment on mask fit factor at the p = .00042 level for the five conditions [F(4, 5) = 44.736, p = .000418]. The heat treatment passed fit testing through 5 treatment cycles, the UVGI treatment failed after one cycle.

There was no significant effect between heat treatment for the two KN95 masks (M = 176.25, SD = 33.78) in comparison to the 2 untreated masks in the control group (M = 178.25, SD = 43.5) on fit factor scores t(2) = 0.078, p = .95.

We did not observe a significant effect for heat treatment for the two 3M 8210 Plus masks (M = 200, SD = 0) compared to the 2 untreated masks in the control group (M = 200, SD = 0) on fit factor scores t(2) = 65535, p = N/A.

The two KN95 masks that received UV treatment (M = 17.75, SD = 3.18) compared to the 2 untreated masks in the control group (M = 178.25, SD = 43.5) demonstrated significantly inferior fit factor scores t(2) = 7.3, p = .02. The fit of the UV treated masks compared to the two KN95 masks that received heat treatment (M = 176.25, SD = 33.78) also produced significantly inferior on fit factor scores t(2) = 11.4, p = .0076.

## RESULTS TABLE

| **GROUPS** | **COUNT** | **SUM** | **AVERAGE** | **VARIANCE** | **POST-HOC P-VALUE** |
| --- | --- | --- | --- | --- | --- |
| KN95 Control | 2 | 356.5 | 178.25 | 946.125 | – |
| KN95 Heat | 2 | 352.5 | 176.25 | 378.125 | .95 |
| KN95 UVGI | 2 | 35.5 | 17.75 | 10.125 | .02 |
| 3M 8210 Control | 2 | 400 | 200 | 0 | – |
| 3M 8210 Heat | 2 | 400 | 200 | 0 | N/A |
| 3M 8210 UVGI | 0 | N/A | N/A | N/A | N/A |
| 8200 Control | 2 | 400 | 200 | 684.5 | – |
| 8200 Heat | 2 | 363 | 181.5 | 0 | .42 |
| 8200 UVGI | 2 | N/A | N/A | N/A | N/A |
| 8511 Control | 2 | 400 | 200 | 0 | – |
| 8511 Heat | 2 | 400 | 200 | 0 | N/A |
| 8511 UVGI | 2 | 259 | 129.5 | 9940.5 | .42 |
| Jackson R20 Control | 2 | 400 | 200 | 0 | – |
| Jackson R20 Heat | 2 | 395 | 197.5 | 12.5 | .42 |
| Jackson R20 UVGI | 2 | 17.6 | 8.8 | 20.48 | .00028 |

## Discussion

The purpose of our study was to assess the fit factor of filtering facepiece respirators (FFR) after two different disinfection methods (75°C Hot Air (30 mins) for 10 cycles) and UVGI = UV 254 nm, 8W, 30 min for 10 cycles. Importantly, we found that while heat did not appreciably degrade the fit factor of masks from two manufacturers, UVGI significantly degraded the fit of all tested models by >90%. The 3M masks were not tested for UVGI due to a shortage of testing materials.

Previous work by Heimbuch et al ^13^ reports testing 15 models but 3 models failed to fit the model head and couldn’t be tested. After 10 cycles of UVGI all tested models indicated a significantly lower peak force for treated straps for at least one condition tested compared to controls and doffing. Nevertheless, Heimbuch et al ^13^ report that UVGI does not significantly degrade fit factor testing after 10 treatment cycles^13^. They report that of six FFR models tested, after 20 treatment cycles, respirators by Kimberly Clark PFR and U.S. Safety AD4N95 failed two of the three fit testing conditions assessed. The untestable mask models in the Heimbuch et al report^13^ were certified by NIOSH and cleared by the FDA. Evaluation of UVGI by Bergman et al^1^. report fit factor was not significantly degraded by treatment of UVGI)=15-min @ 254nm, however they did not test the straps and only one side of the N95 faced the lamp. The 3M company^7^ tested N95 masks UVGI 30- min @ 254nm (15-min per side) and they report elasticity loss, a strong burnt odor and nose foam compression on some models. Straps are an unescapable component of respiratory mask fit.

We note that in these previous studies, testing was performed only on static mannequins using three fitting tests, compared to our use of a human model and the OSHA fit testing protocol that utilizes eight fitting tests including dynamic maneuvers such as turning the head side-to-side, up-and-down, talking, and bending over. These dynamic fitting maneuvers could have revealed fitting inadequacies caused by UVGI that were not present on static mannequin testing.

## LIMITATIONS OF OUR STUDY

Due to a US supply chain shortage of N95 masks, our design was limited in the ability to study a variety of disinfecting conditions. One limitation of this study is our comparison of five cycles of hot air treatment to ten cycles of UVGI. Our goal in this design was to err on the minimum viable number of cycles of heat treatment in this first pilot study to detect adequate fit factor after heat treatment, given the emergent nature of the US PPE shortage and need for fit factor validation of this unstudied disinfection method. Because others have already validated UVGI for 10 and 20 cycles and previously published their validity in preserving fit factor^13^, we felt comfortable in setting 10 cycles of UVGI for our study to allow direct comparison to prior work.

This research needs to be replicated with a larger number of masks using the brands most commonly used by clinicians. Due to a shortage of 3M 8210 N95 masks, we were not able to obtain sufficient samples for UVGI testing in this study. We were unable to test the 3M 8210 for UVGI as the mask shortage meant the 4c lab was unable to attain these masks for testing.

Therefore, another limitation of this current study is that of only testing UV treatment on the mask materials from 3 manufacturers and 5 models. Tsai 2020^14^, the N95 inventor, and Lindsley 2015^15^ note that UV damage to polypropylene fibers is possible but that it may be dosage, model and frequency dependent which is consistent with our preliminary disinfection and fit data. As we have previously noted, other studies using mannequins have studied a larger number of materials from a diversity of mask manufacturers.

## Conclusion

Our research is concordant with previous research in our observation that we observed mask models that failed after UVGI treatment using various fit testing methods. These data suggest that UVGI methods of FFR decontamination cause fit failure in more than 40% of the models tested to date. FFR fit testing may be needed after each decontamination cycle for each user to confirm proper fit in order to ensure that decontaminated masks perform according to CDC guidance that “[a] key consideration for safe extended use is that the respirator must maintain its fit and function.”

## FUTURE WORK

There is evidence to show that donning and doffing frequency^16^alters fit and is a frequent source of cross contamination and self-inoculation^8^. Future testing by our group will show the effect of disinfecting mask models subjected to extended use. There are four areas which merit further consideration. Will (75°C Hot Air (30 mins) for 10 cycles) and UVGI = UV (254 nm, 8W, (30 min) for 10 cycles) decontamination harm the fit of other N95 mask models? Can the method of decontamination work through all the layers of trapped virus in the particles? Will turning the mask as required in UVGI testing expose the cleaner or user to additional infection risk? These tests need to be performed with the Covid19 virus. Our group and other excellent and resourceful scientists around the world are addressing these important and pressing questions.

This article is for information purposes only. We cannot make any recommendations or provide any guidance on the possible disinfection and reuse of respirators or other PPE. Here is a link to the current NIOSH guidance on Strategies for Optimizing the Supply of N95 Respirators^17^: https://www.cdc.gov/coronavirus/2019-ncov/hcp/respirators-strategy/index.html

We do not advocate or advise on practices and suggest that you strictly adhere to your hospital’s policies and procedures. Our goal is to provide the best data available at the time of publication.

## Data Availability

Data are available upon request to the corresponding author

